# At the head and heart of oxytocin’s stress-regulatory neural and cardiac effects: a chronic administration RCT in children with autism

**DOI:** 10.1101/2023.04.04.23288109

**Authors:** Kaat Alaerts, Nicky Daniels, Matthijs Moerkerke, Margaux Evenepoel, Tiffany Tang, Stephanie Van der Donck, Viktoria Chubar, Stephan Claes, Jean Steyaert, Bart Boets, Jellina Prinsen

## Abstract

Intranasal administration of oxytocin is increasingly explored as a new approach to facilitate social development and reduce disability associated with a diagnosis of autism spectrum disorder (ASD). In light of the growing number of trials, it is crucial to gain deeper insights into the neuroplastic changes that are induced from multiple-dose, chronic use of oxytocin, over a course of weeks. To date however, oxytocin’s chronic neuromodulatory impact in the pediatric brain remains unknown.

Here, we present a double-blind, randomized, placebo-controlled pharmaco-neuroimaging trial examining the neural effects of a four-week intranasal oxytocin administration regime (12 IU, twice daily) in pre-pubertal school-aged children with ASD (8-12 years, 45 boys, 12 girls). Resting-state fMRI scanning and simultaneous, in-scanner heart rate measurements were assessed before, immediately after and four weeks after the nasal spray administration period.

Four weeks of chronic oxytocin administration in children with ASD induced significant reductions in intrinsic functional connectivity between amygdala and orbitofrontal cortex, particularly at the four-week follow-up session, thereby replicating prior observations of neuromodulatory changes in the adult brain. Notably, the observed reductions in amygdala-orbitofrontal connectivity were associated with improved autonomic stress-regulation, indexed by increased high-frequency heart rate variability. Further, oxytocin’s neural and cardiac autonomic effects were significantly modulated by epigenetic modifications of the oxytocin receptor gene, indicating that oxytocin-induced stress-regulatory effects were more pronounced in children with reduced epigenetic methylation, and thus higher oxytocin receptor expression. Finally, whole-brain exploratory functional connectivity analyses also revealed an overall oxytocin-induced enhancing effect on amygdala coupling to regions of the salience network (insula, anterior cingulate cortex), likely reflective of oxytocin’s (social) salience effects.

Together, these observations provide initial insights into the stress-regulatory neural and cardiac effects induced by chronic oxytocin administration in children with ASD, and point toward important epigenetic modulators that may explain inter-individual variations in oxytocin-induced responses.

## Introduction

Intranasal administration of the neuropeptide oxytocin is increasingly considered as a novel approach for alleviating disabilities associated with an autism spectrum disorder (ASD) diagnosis, including difficulties in social communication and interaction, and restricted and repetitive behaviors and interests ^1^.

In relation to ASD, initial single-dose oxytocin administration studies consistently demonstrated behavioral improvements on various social tasks, including tasks assessing affective speech comprehension (emotional intonations) ^2, 3^, emotion recognition ^4, 5^, repetitive behavior ^6^ and social decision making (cyberball computer game) ^7^ (for review, see e.g., ^8^). Recent multiple-dose, chronic administration studies have yielded a more mixed pattern of effects, with some studies showing beneficial clinical effects ^9-15^, while others identified no benefit of oxytocin over placebo nasal spray administration ^16-19^.

In light of this mixed pattern of effects, it is crucial to gain a deeper understanding of the neural substrates that underlie behavioral effects, allowing to delineate possible mechanisms of inter-individual variation in clinical treatment responses. To date, insights into the neural effects of oxytocin have predominantly emerged from pharmaco-neuroimaging studies examining the acute effects of single-dose administrations in adults, predominantly highlighting a key role for the amygdala in exerting oxytocin’s neuromodulatory action ^20-22^. Specifically, prior single-dose administration studies in neurotypical and clinical populations (including ASD) generally demonstrated an attenuating effect of acute oxytocin administration on task-evoked amygdala reactivity, presumed to reflect oxytocin’s arousal-dampening and anxiolytic effects ^20, 23^. In particular, a seminal study by Kirsch et al. (2005) showed that a single-dose of oxytocin elicits an acute attenuating effect on amygdala reactivity and amygdala–brainstem connectivity ^24^, suggestive of oxytocin’s impact on regulating autonomic arousal, sympathetic-parasympathetic tone and fear behavior. However, a more complex pattern of results emerged from subsequent resting-state fMRI studies, with some showing an acute increase in amygdala functional connectivity to prefrontal regions, while others showed reductions in amygdala coupling or no effects after a single-dose administration (for a recent review see ^21^).

Insights in how repeated, daily administrations of oxytocin affect neural circuits are still highly sparse, with only two pharmaco-neuroimaging studies addressing this topic, both in adults with ASD. In a first study, Watanabe et al. (2015) explored the neural effects of a six-week oxytocin administration regime in 17 adult men with ASD and reported an increase in connectivity between the anterior cingulate and prefrontal cortex ^10^. Note however, that the reported multiple-dose, chronic effect may - at least in part - reflect an acute effect of exogenously administered oxytocin, considering that participants were scanned approximately 15 or 40 min after the last nasal spray administration (of the six-week regime), which corresponds to the optimal time frame for assessing acute, single-dose effects. In another study, 38 adult men with ASD received oxytocin or placebo nasal spray, daily for a period of four weeks and chronic effects of the repeated administration were examined at least 24 hours after the last nasal administration. In the latter study, chronic oxytocin administration elicited an attenuation of amygdala reactivity, both intrinsically during resting-state scanning ^25^ and task-evoked ^26^, further highlighting a predominant anxiolytic impact also after chronic oxytocin administration. Chronic oxytocin-induced changes in intrinsic functional connectivity of amygdala regions were also observed, predominantly indicating reduced coupling of the amygdala to prefrontal and orbitofrontal cortices, which was interpreted to reflect a reduced need to downregulate amygdala activity via frontal top-down control ^27^. Importantly, neural adaptations in amygdala-orbitofrontal connectivity were shown to outlast the period of actual administration until one month and even one year after the nasal spray administration period, and were associated with behavioral improvements in repetitive behavior and attachment style ^26, 27^. These observations thereby provided important evidence that in the adult brain, chronic oxytocin administration can induce long-lasting neuroplastic changes in amygdala circuitry that associate to retained clinical improvements.

However, a more comprehensive and integrative understanding of the neuromodulatory impact of chronic oxytocin administration is needed, especially for pediatric populations. This is of particular importance, considering that ASD is a neurodevelopmental condition for which therapeutic approaches can be preferably administered in early life developmental windows ^28^. To fill this gap, here, we present a first pharmaco-neuroimaging study with school-aged boys and girls with ASD, examining the neural effects of a four-week course of chronic oxytocin administration on amygdala functional connectivity as assessed using resting-state fMRI scanning. To examine whether chronic oxytocin administration could also induce long-lasting neuroplastic changes in the pediatric brain, we also included a follow-up resting-state fMRI assessment four weeks after cessation of the daily administrations. Primary analyses examined whether oxytocin administration would induce similar attenuating effects on amygdala-orbitofrontal connectivity in school-aged boys and girls with ASD, as previously identified in adult men with ASD ^27^. Additionally, exploratory analyses were performed, to obtain a complete picture of how chronic oxytocin impacts whole-brain connectivity of amygdala regions.

To obtain a more integrative assessment of oxytocin’s purported anxiolytic role in modulating cardiac autonomic arousal and sympathetic-parasympathetic homeostatic balance, we additionally performed in-scanner assessments of heart rate variability (HRV), an established marker of cardiac autonomic nervous system (ANS) activity ^29^. The high-frequency component of HRV uniquely represents the contribution of the parasympathetic system via the nervus vagus and is generally regarded to constitute an important marker of psychophysiological homeostasis and well-being ^29^. While insights into the effect of oxytocin on HRV are still relatively sparse, existing single-dose administration studies demonstrated acute effects on facilitating an increase in resting-state high-frequency HRV ^30-34^. In this study, we examined, for the first time, whether chronic oxytocin administrations can elicit similar cardiac autonomic effects, and perhaps a (long-lasting) retention of heightened intrinsic cardiac parasympathetic tone.

Finally, there is a growing awareness that oxytocin nasal spray administration effects may not be uniform and that distinct context- and/or person-dependent factors may hamper or facilitate treatment responses ^35^. Since oxytocin’s effects on brain function and behavior are known to be mediated by the prevalence of oxytocin receptors as determined by the oxytocin receptor gene (*OXTR*), it is increasingly put forward that (epigenetic) variations of *OXTR* may form an important factor for explaining variable treatment responses ^36-38^, but empirical evidence is largely lacking. To investigate the impact of *OXTR* epigenetics, we examined whether variability in neural effects is mediated by variability in baseline (pre-nasal spray) DNA methylation of the *OXTR* gene.

## Methods

### General study design

This double-blind, randomized placebo-controlled trial with between-subject design investigated the neural effects of chronic oxytocin administration on intrinsic functional connectivity of the amygdala in school-aged children with ASD. Resting-state fMRI scanning and HRV assessments were performed at baseline (T0); immediately after four consecutive weeks of daily nasal spray administrations (24 hours after the last nasal spray administration) (T1); and at a follow-up session, four weeks after cessation of the nasal spray administration period (T2).

Please see **Figure 1** for the CONSORT Flow diagram visualizing the number of participants randomized and analyzed. Please also see **Supplementary Methods** outlining the impact of COVID-19 related health restrictions on the recruitment and flow of participants in the trial.

**Figure 1.**
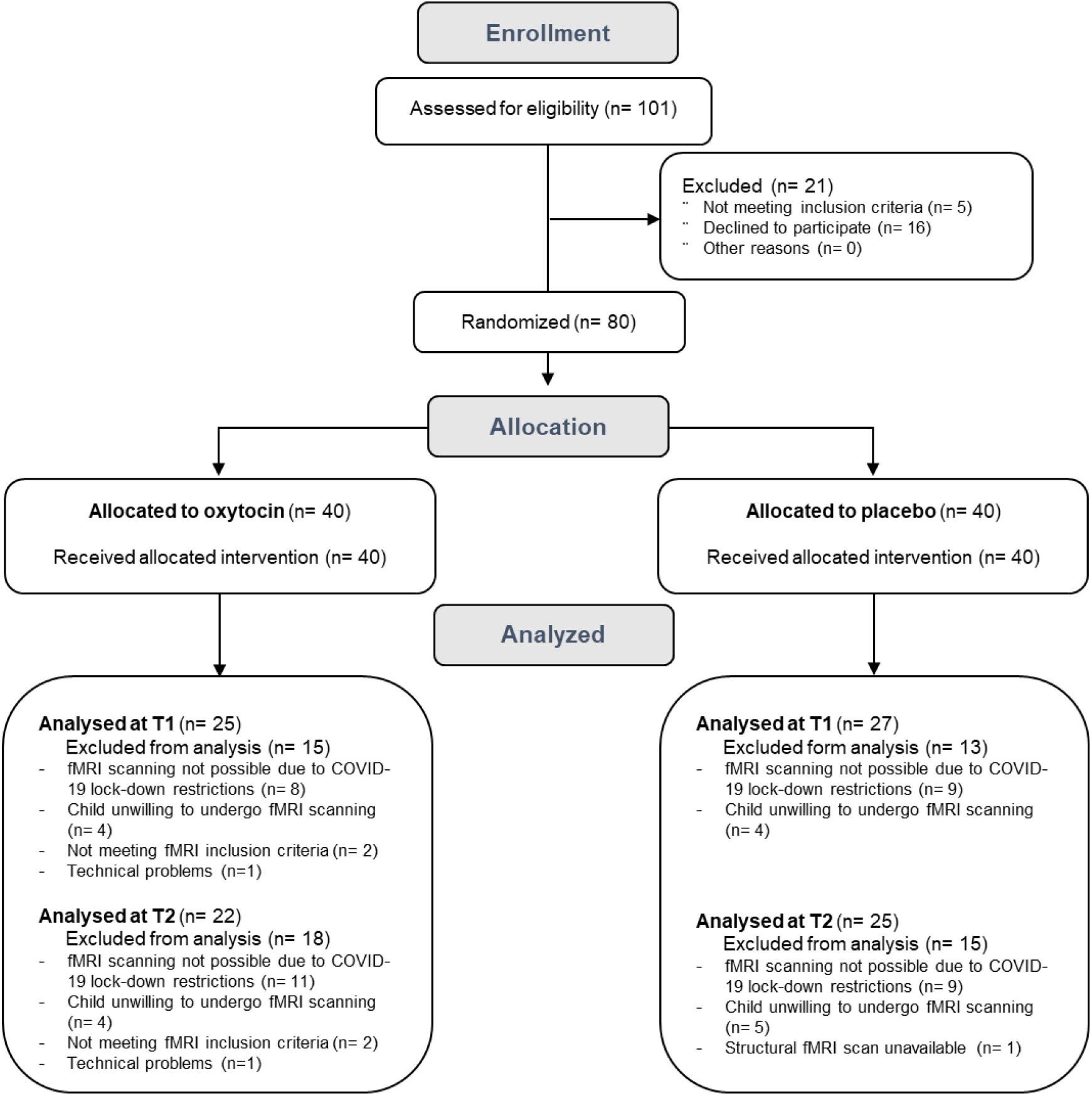
CONSORT flow diagram of participants in the trial. MRI scanning was performed at baseline (T0), after the four-week (oxytocin/placebo) nasal spray administration period (T1), and at a follow-up session, four weeks after cessation of the nasal spray administration period (T2).

Written informed consent from the parents and assent from the child were obtained prior to the study. Consent forms and study design were approved by the local Ethics Committee for Biomedical Research at the University of Leuven, KU Leuven (S61358) in accordance with The Code of Ethics of the World Medical Association (Declaration of Helsinki). The neural assessments were conducted at the Leuven University Hospital (registered at the EU Clinical Trials register: EudraCT 2018-000769-35 and the Belgian Federal Agency for Medicines and Health products). As indicated in the EudraCT registration, fMRI data collections were part of a broader assessment including behavioral-clinical characterizations ^19^, as well as other (neuro)physiological (electroencephalographic) and biological assessments (reports in preparation). The trial was monitored by the Clinical Trial Center at the University hospital of Leuven, and all trial staff had Good Clinical Practice certification and was trained in the study protocol.

### Participants

Children with a formal diagnosis of ASD were recruited through the Autism Expertise Centre at the Leuven University Hospital between July 2019 and January 2021 (**Figure 1, Table 1**). The diagnosis was established by a multidisciplinary neuropediatric team based on the strict criteria of the DSM-5 (Diagnostic and Statistical Manual of Mental Disorders) ^1^. Prior to randomization, the Autism Diagnostic Observation Schedule (ADOS-2) ^39^ and estimates of intelligence (four subtests of the Wechsler Intelligence Scale for Children, Fifth Edition, Dutch version) ^40^ were acquired (**Table 1**).

**Table 1.**
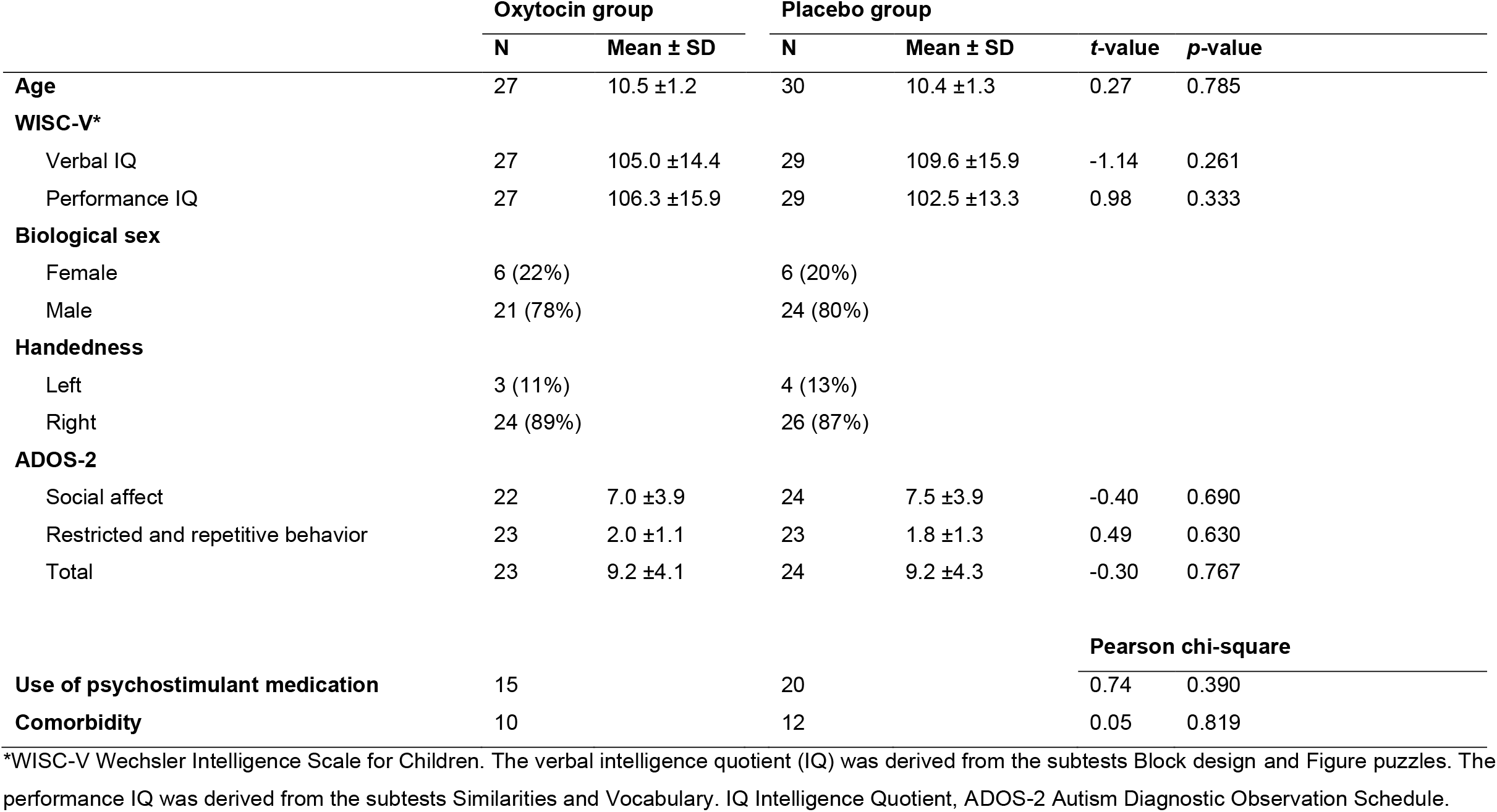
Demographic and clinical characteristics of children randomized to receive oxytocin or placebo. Mean baseline scores are listed separately for each nasal spray administration group. T- and p-values correspond to independent sample t-tests assessing between-group differences in baseline scores.

|  | Oxytocin group |  | Placebo group |  | t-value | p-value |
| --- | --- | --- | --- | --- | --- | --- |
| | N | Mean ± SD | N | Mean ± SD | | |
| **Age** | 27 | 10.5 ± 1.2 | 30 | 10.4 ± 1.3 | 0.27 | 0.785 |
| **WISC-V\*** |  |  |  |  |  |  |
| Verbal IQ | 27 | 105.0 ± 14.4 | 29 | 109.6 ± 15.9 | -1.14 | 0.261 |
| Performance IQ | 27 | 106.3 ± 15.9 | 29 | 102.5 ± 13.3 | 0.98 | 0.333 |
| **Biological sex** |  |  |  |  |  |  |
| Female | 6 (22%) |  | 6 (20%) |  |  |  |
| Male | 21 (78%) |  | 24 (80%) |  |  |  |
| **Handedness** |  |  |  |  |  |  |
| Left | 3 (11%) |  | 4 (13%) |  |  |  |
| Right | 24 (89%) |  | 26 (87%) |  |  |  |
| **ADOS-2** |  |  |  |  |  |  |
| Social affect | 22 | 7.0 ± 3.9 | 24 | 7.5 ± 3.9 | -0.40 | 0.690 |
| Restricted and repetitive behavior | 23 | 2.0 ± 1.1 | 23 | 1.8 ± 1.3 | 0.49 | 0.630 |
| Total | 23 | 9.2 ± 4.1 | 24 | 9.2 ± 4.3 | -0.30 | 0.767 |
|  |  |  |  |  | **Pearson chi-square** |  |
| **Use of psychostimulant medication** | 15 |  | 20 |  | 0.74 | 0.390 |
| **Comorbidity** | 10 |  | 12 |  | 0.05 | 0.819 |

\*WISC-V Wechsler Intelligence Scale for Children. The verbal intelligence quotient (IQ) was derived from the subtests Block design and Figure puzzles. The performance IQ was derived from the subtests Similarities and Vocabulary. IQ Intelligence Quotient, ADOS-2 Autism Diagnostic Observation Schedule.

Principal inclusion criteria comprised a clinical diagnosis of ASD, age (8-12 years old), intelligence quotient (IQ) above 70, native Dutch speaker, a stable background treatment for at least four weeks prior to the screening and no anticipated changes during the trial. Only premenstrual girls were included. Principal criteria for exclusion comprised any neurological (e.g., stroke, epilepsy, concussion), significant physical disorder (liver, renal, cardiac pathology), prior use of oxytocin, or any contraindication for magnetic resonance imaging (MRI). The presence of comorbid psychiatric disorders and current psychoactive medication use was screened for and logged (see **Table 1** and ^19^).

Since this is the first pharmaco-neuroimaging study examining the neural effects of chronic oxytocin administration in children with ASD, formal power calculation based on existing literature was difficult. In a prior study from our lab with a similar parallel clinical trial design in adults with ASD ^27^, four weeks of chronic oxytocin versus placebo nasal spray administration yielded a reduction in amygdala-prefrontal functional connectivity with an overall effect size of *d* = 1.057. Sample sizes for the functional connectivity analyses in the current study were therefore set at 50 participants (25 oxytocin, 25 placebo) allowing to detect a similar effect size with α = 0.05 and 90% power. Thus far, no prior studies investigated the effect of chronic oxytocin administration on resting-state HRV.

### Study medication and dosing

Participants were randomized (permuted-block randomization) to receive oxytocin (Syntocinon®, Sigma-tau) or placebo nasal sprays, administered in identical blinded amber 10 ml glass bottles with metered pump (preparation, packaging, blinding by Heidelberg University Hospital, Germany). The placebo spray consisted of all the ingredients used in the active solution except the oxytocin compound. Participants were randomly assigned in a 1:1 ratio, with stratification according to age, IQ and biological sex. All research staff conducting the trial, participants and their parents were blinded to nasal spray allocation.

Children (assisted by their parents) were asked to self-administer a daily dose of 2 × 12 international units (IU) nasal spray or placebo equivalent (3 puffs of 2 IU in each nostril), 12 IU in the morning and 12 IU in the afternoon, during 28 consecutive days. Participants received clear instructions about use of the nasal sprays ^41^ through a demonstration together with the experimenter. More information regarding nasal spray adherence and side effects screening is provided in **Supplementary Methods**.

### MRI data acquisition and functional connectivity analysis

#### MRI data acquisition

Anatomical and resting-state fMRI images (7 min, eyes open) were acquired on a 3.0 Tesla Philips MR scanner (Best, The Netherlands) with a 32-channel phased-array head coil. Detailed information on the scanning parameters, MRI data pre-processing and in-scanner head motion analyses are provided in **Supplementary Methods**.

#### Amygdala-orbitofrontal connectivity

Region-of-interest (ROI) connectivity analysis was performed assessing nasal spray-induced changes in connectivity between bilateral amygdala and orbitofrontal cortex (OFC) (see **Figure 2A**). The ROIs were identical to those adopted in a prior chronic oxytocin fMRI pharmaco-neuroimaging study in adult men with ASD ^27^, as defined by the FSL Harvard-Oxford subcortical and cortical atlas. As implemented in the CONN functional connectivity toolbox version 17.f ^42^, mean time-series were extracted by averaging across all voxels in each ROI and bivariate correlation coefficients were computed between the time-course of the amygdala seeds (left and right) and the time-courses of the left and right orbitofrontal ROIs. Correlation values were Fisher z-transformed. In addition to the hypothesis-driven ROI analysis, exploratory whole-brain analyses were performed by assessing nasal spray-induced changes in functional connectivity between bilateral amygdala (seeds) and all other regions of the cortical (n = 91) and subcortical (n = 13) Harvard-Oxford atlas.

**Figure 2.**
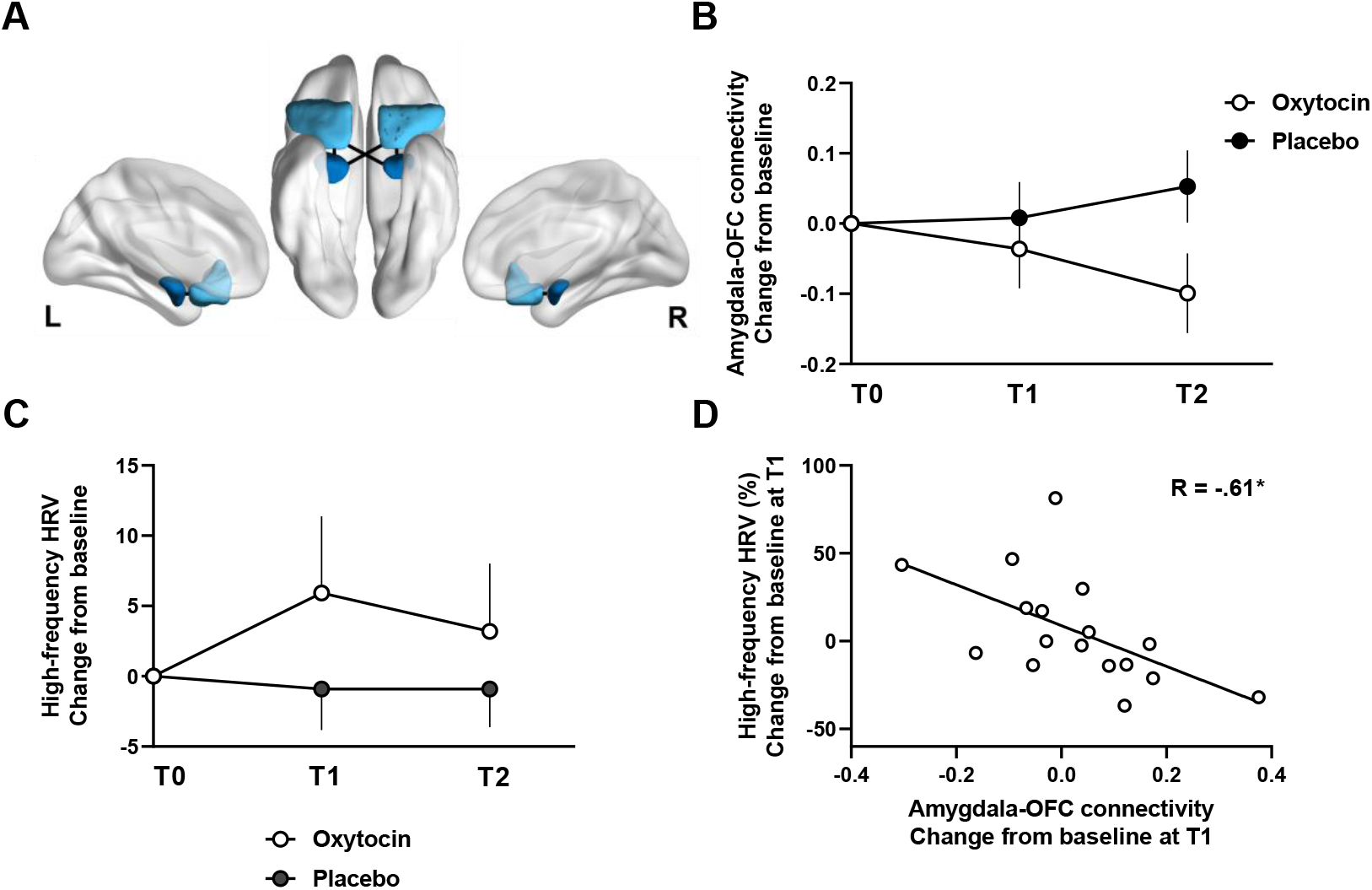
Oxytocin-induced changes in amygdala-orbitofrontal connectivity and HRV. **Panel A** visualizes regions of interest (ROIs) in bilateral amygdala (dark blue) and orbitofrontal cortex (light blue), as defined by the FSL Harvard-Oxford subcortical and cortical atlas. ROIs are overlaid on inflated surface maps generated using BrainNet Viewer (http://www.nitrc.org/projects/bnv/). **Panel B** visualizes changes from baseline in amygdala-orbitofrontal connectivity separately for each nasal spray group (oxytocin and placebo) and assessment session (T1–T2). **Panel C** visualize changes from baseline in high-frequency HRV separately for each nasal spray group (oxytocin and placebo) and assessment session (T1–T2). **Panel D** visualizes the relationship between oxytocin-induced changes in amygdala-orbitofrontal functional connectivity and oxytocin-induced changes in high-frequency HRV at T1. Vertical bars denote ±standard errors. T1: assessment session immediately after the four-week nasal spray administration period (at least 24 h after the final administration); T2: assessment session four weeks after cessation of the nasal spray administrations. OFC: orbitofrontal cortex. R: Spearman correlation coefficient. * denotes p < .05.

### Heart rate variability recordings and data handling

Photoplethysmography (PPG) recordings were performed simultaneous to the resting-state fMRI recording by means of a wireless Peripheral Pulse Unit MRI Sensor (Philips) (sampling rate: 500 Hz). The PPG sensor was placed over the index finger of the non-dominant hand to monitor blood volume changes in the microvascular bed of the underlying tissue. The time intervals between blood volume pulse waves were assessed using Kubios HRV Premium software (version 3.5.0) ^43^ to derive continuous inter-beat-intervals (IBI) for assessing heart rate variability (HRV). All IBI time series were manually inspected prior to analysis and automatic artifact removal, as implemented in Kubios, was performed. At T1, PPG recordings were available for 43 participants (20 oxytocin/23 placebo) and for 40 participants (18 oxytocin/22 placebo) at T2. Upon artefact removal, 4 additional participants (3 oxytocin/1 placebo) were removed from the final analyses (availability of less than 5 min of noise-free data and/or >5% ectopic beats ^44^). For each of the remaining subjects, HRV frequency domain analyses were performed using Fast Fourier Transformation based on Welch’s periodogram. Percentage power values were computed for the high-frequency component of the HRV signal (HF-HRV%), which in children is defined between 0.24 – 1.04 Hz ^45^.

### Assessment of baseline DNA methylation of the oxytocin receptor gene

Salivary samples were collected from each child at the end of their baseline (T0) study visit using the Oragene DNA sample collection kit (DNA Genotek Inc., Canada), to explore epigenetic variations of the oxytocin receptor gene (*OXTR)*. An important epigenetic mechanism is the methylation of a cytosine from a CpG dinucleotide (i.e. a cytosine followed by a guanine, connected by a phosphate) within the DNA, which is generally associated with a silencing of the gene and thus a decrease in gene transcription ^46^. Here, DNA methylation was assessed at three CpG sites of *OXTR* that have been shown to be impacted in autism (i.e., -934, -924 and -914; see ^46^). More detailed information regarding the DNA collection procedures and analyses are provided in **Supplementary methods**.

### Statistical analysis

To assess oxytocin-induced effects, amygdala-orbitofrontal functional connectivity z-transformed r-values were subjected to mixed-effects analyses of variances. The factor ‘subject’ was inserted as a random effect and the factors ‘nasal spray’ (oxytocin, placebo), ‘session’ (T1, T2), ‘amygdala seed’ (left, right) and ‘OFC ROI’ (left, right), as well as all interactions with the factor ‘nasal spray’, were inserted as fixed effects. To correct for variance in the individuals’ baseline T0 scores, baseline values prior to nasal administration were included as a covariate in the model.

Oxytocin-induced effects in high-frequency HRV were analyzed using similar mixed models, i.e., with the factors ‘nasal spray’ (oxytocin, placebo) and ‘session’ (T1, T2), and baseline (T0) scores modeled as covariate.

Spearman correlation analyses were performed to examine possible associations between neural and cardiac autonomic changes observed in the oxytocin group. Spearman correlations were also performed between (baseline) OXTR DNA methylation (average across CpG sites) and oxytocin-induced changes in amygdala-orbitofrontal connectivity and high-frequency HRV to examine possible moderations of neural and cardiac oxytocin-induced effects by variations in epigenetic modifications of the oxytocin receptor gene.

Additionally, exploratory, oxytocin-induced changes in whole-brain amygdala connectivity analyses were performed separately for each assessment session (T1, T2) by subjecting change from baseline connectivity scores to independent t-tests with the between-group factor ‘nasal spray’ (oxytocin, placebo). Considering the exploratory nature of the whole-brain analysis, amygdala connections displaying significant oxytocin-induced effects are reported at an uncorrected *p* < .05 threshold (connection-level, CONN-toolbox).

### Data availability

The data that support the findings of this study are available on request from the corresponding author, KA.

## Results

### Oxytocin-induced changes in amygdala-orbitofrontal connectivity

Mixed-effects analyses revealed a main effect of ‘nasal spray’, indicating an attenuation in amygdala-orbitofrontal connectivity in the oxytocin group, compared to the placebo group (*F*(1, 52.97) = 4.26; *p* = .044; *ŋ*^*2*^ = .07) (**Figure 2B**). Also a ‘treatment x session’ interaction effect was evident (*F*(1, 305) = 7.26; *p* = .007; *ŋ*^*2*^ = .02), indicating that the oxytocin-induced attenuation in amygdala-orbitofrontal connectivity was evident at the four-week follow-up session (T2, Bonferroni post-hoc, *p* < .001), not at the T1 post-session (*p* > .100) (**Figure 2B**). None of the other main or interaction effects reached significance (all, *p* > .05).

### Oxytocin-induced changes in high-frequency HRV

Mixed-effect analysis of high-frequency HRV yielded a main effect of ‘treatment’ (*F*(1, 76) = 4.67; *p* = .034; *ŋ*^*2*^ = .057), indicating overall higher parasympathetic high-frequency HRV in the oxytocin, compared to the placebo group (**Figure 2C**). The main effect of ‘session’ or the ‘treatment x session’ interaction were not significant (both, *p* > .05).

### Association between neural and cardiac autonomic oxytocin-induced changes

Spearman correlation analyses revealed a significant association between neural changes in amygdala-orbitofrontal connectivity and cardiac autonomic changes in high-frequency HRV, indicating that children who display a stronger oxytocin-induced attenuation in amygdala-orbitofrontal connectivity at the T1 assessment session, displayed a stronger oxytocin-induced increase in high-frequency HRV (Spearman *ρ* = -.61; *p* = .009) (**Figure 2D**). No significant associations were evident at the T2 follow-up session (*p* > .05).

### Modulation of oxytocin-induced responses by OXTR epigenetic variations

Baseline variations in *OXTR* DNA methylation averaged across CpG sites -934, -924 and -914 were predictive of oxytocin-induced changes in amygdala-orbitofrontal connectivity, indicating that children with low *OXTR* DNA methylation (associated with higher *OXTR* expression) displayed a stronger effect of chronic oxytocin administration on attenuating amygdala-orbitofrontal connectivity, immediately after the nasal spray administration period, at T1 (Spearman *ρ* = .50; *p* = .011) (**Figure 3A**). The retention of oxytocin-induced neural changes at T2 was not significantly modulated by variations in *OXTR* DNA methylation (*p* > .05) (**Figure 3B**).

**Figure 3.**
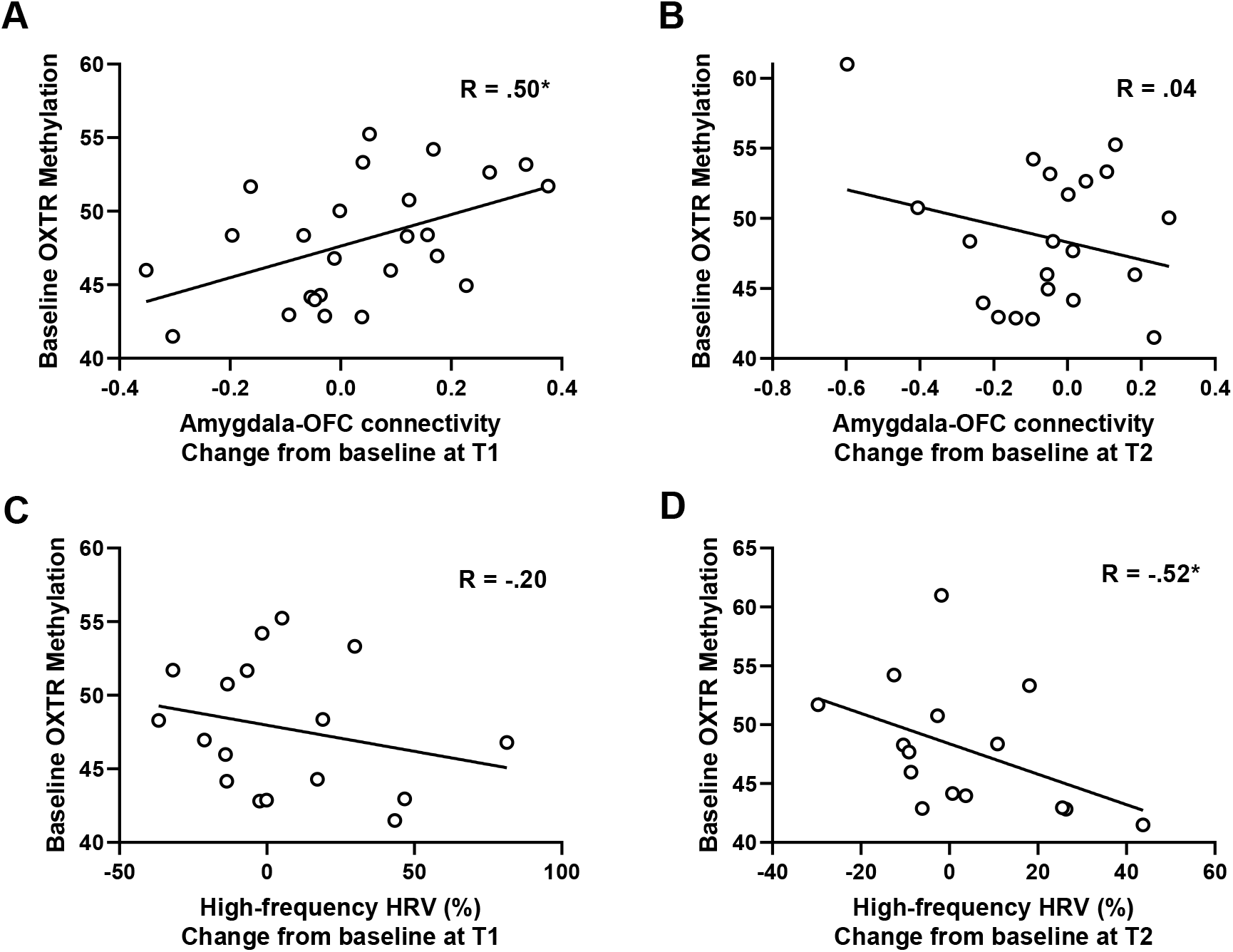
Modulation of oxytocin-induced neural and cardiac effects by *OXTR* epigenetic variations. In terms of functional connectivity, a significant association was identified between baseline variations in *OXTR* methylation of CpG sites -934, -924 and -914 (average) and oxytocin-induced changes in amygdala-orbitofrontal functional connectivity at T1 (**panel A**), not at T2 (**panel B**). In terms of HRV, a significant association was identified between baseline variations in *OXTR* methylation and oxytocin-induced changes in high-frequency HRV at retention session T2 (**panel D**), not at T1 (**panel C**). R: Spearman correlation coefficient. * denotes p < .05.

Modulations of oxytocin-induced changes in high-frequency HRV on the other hand, were significant for the T2 retention session (not for the T1 session), indicating that children with low *OXTR* DNA methylation frequencies displayed a stronger retention of increased high-frequency HRV (Spearman *ρ* = -.52; *p* = .046) (**Figure 3C, D**).

### Exploratory whole-brain amygdala functional connectivity analyses

In addition to the hypothesis-driven exploration of oxytocin’s effects on amygdala-orbitofrontal cortex connectivity, we also performed exploratory whole-brain analyses examining oxytocin-induced changes in functional connectivity between bilateral amygdala (seeds) and all other regions of the cortical (*n* = 91) and subcortical (*n* = 13) Harvard-Oxford Atlas.

At the T1 post session, an overall pattern of enhanced amygdala connectivity was identified (**Figure 4, Supplementary Table 1**), particularly to regions of the salience network, including bilateral insular cortex (including posterior insula in planum polare regions), the anterior cingulate cortex and right supplementary motor area. Increased amygdala connectivity was also evident to cortical regions in frontal, parietal and temporal regions overlapping with the action observation network, i.e., including bilateral inferior frontal gyrus, pars triangularis and pars opercularis, and right supramarginal gyrus (encompassing inferior parietal lobule) and middle temporal gyrus (overlapping with the superior temporal sulcus). Increased subcortical amygdala connectivity was also evident with the globus pallidum.

**Figure 4.**
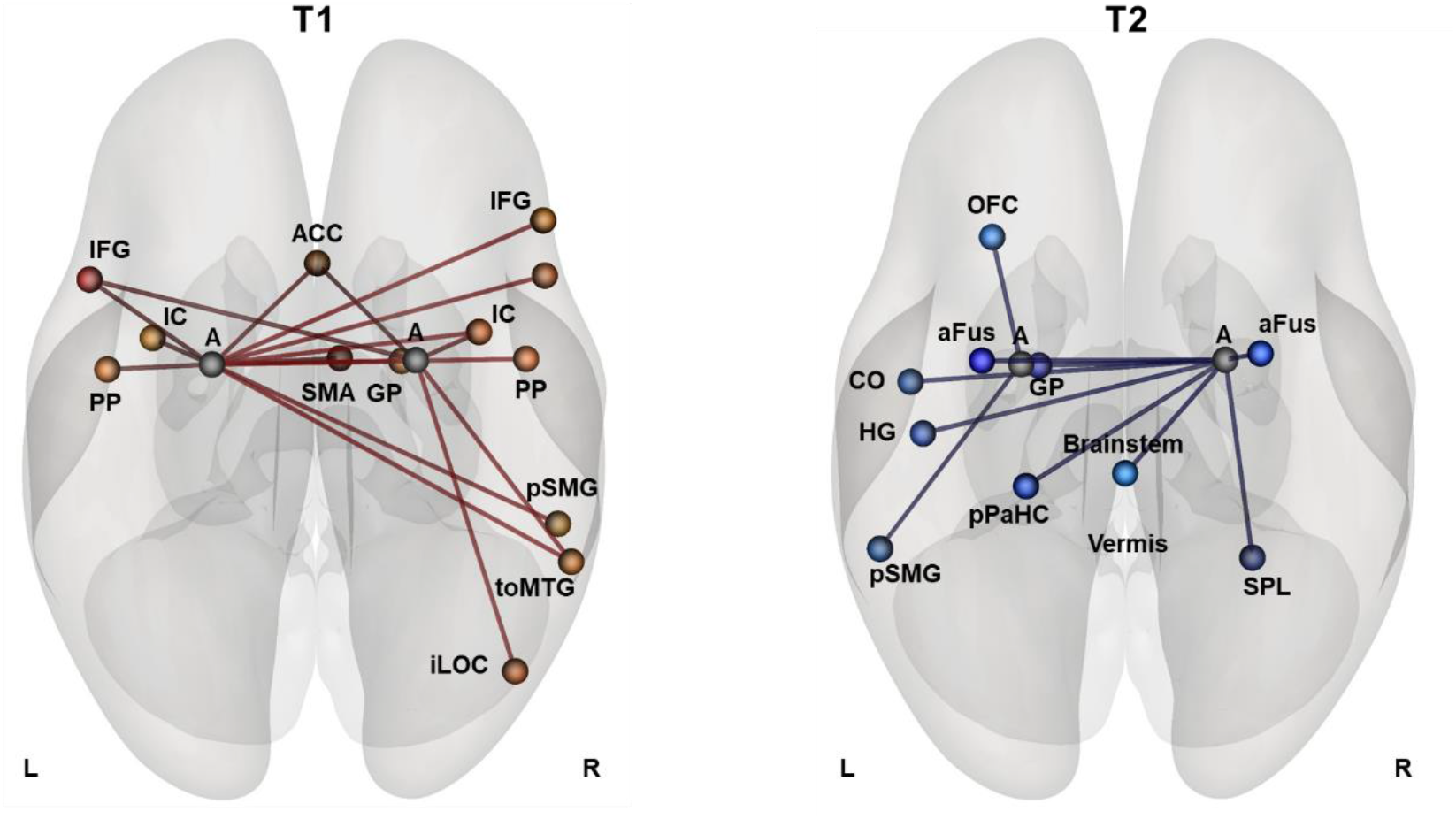
Oxytocin-induced changes in whole-brain amygdala connectivity. Whole-brain analyses exploring oxytocin-induced changes in amygdala connectivity to regions of the cortical (*n* = 91), and subcortical (*n* = 13) Harvard-Oxford Atlas revealed an overall pattern of oxytocin-induced *increases* in amygdala connectivity at the post assessment session T1, and an overall pattern of decreased amygdala connectivity at the four-week follow-up session T2. Connections displaying significant oxytocin-induced effects (blue connections: oxytocin < placebo; red connections: oxytocin > placebo) are reported separately for each assessment session (T1, T2) at an uncorrected *p* <.05 threshold. T1: assessment session immediately after the four-week nasal spray administration period (at least 24 hours after the final administration); T2: assessment session four weeks after cessation of the nasal spray administrations. A: Amygdala; ACC: Anterior Cingulate Cortex; aFus: anterior Fusiform Gyrus; CO: Central Opercular Cortex; GP: Globus Pallidus; HG: Heschle’s Gyrus; IC: Insular Cortex; IFG: Inferior Frontal Gyrus; iLOC: inferior Lateral Occipital Cortex; OFC: Orbitofrontal Cortex; PP: Planum Polare; pPaHC: posterior Parahippocampal cortex; pSMG: posterior Supramarginal Gyrus; SMA: Supplementary Motor Area; SPL: Superior Parietal Lobule; L: left; R: right. toMTG: temporo-occipital Middle Temporal Gyrus;

At the T2 follow-up session, four weeks after cessation of the nasal spray administrations, a remarkably different pattern of oxytocin-induced changes in amygdala connectivity was evident, indicating an emergence of reduced amygdala connectivity to distinct cortical and subcortical regions (**Figure 4, Supplementary Table 1**). Particularly, in addition to reductions in amygdala-orbitofrontal connectivity, reductions in amygdala connectivity were identified with bilateral temporal fusiform cortex, right transverse temporal gyrus (Heschle’s gyrus), opercular cortex and parietal regions including the supramarginal gyrus and superior parietal lobule. Reductions in amygdala-subcortical connectivity were mainly identified for the brainstem, the globus pallidum and parahippocampal structures.

## Discussion

In this pharmaco-neuroimaging study examining the neural effect of chronic oxytocin administration in children with ASD, we identified significant changes in intrinsic functional connectivity of the amygdala up to four weeks after cessation of the actual nasal spray administration period.

Prior observations of neuroplastic changes in intrinsic functional connectivity between amygdala and orbitofrontal cortex, as seen in the autistic male adult brain ^27^, were replicated in the current pediatric population of boys and girls with autism, highlighting the importance of this amygdala-centered circuit in oxytocin’s neuromodulatory effects. In children, however, the pattern of reduced amygdala-orbitofrontal connectivity only emerged significantly at the follow-up session, four weeks after cessation of the daily administrations, whereas in the adult sample, the effect already emerged significantly immediately after the nasal spray administration period and further enlarged at follow-up.

Generally, adaptations in amygdala-prefrontal/orbitofrontal interactions are interpreted to reflect changes in top-down inhibitory control of prefrontal regions over amygdala reactivity ^47^. However, considering that the repeated oxytocin administrations may have halted excessive amygdala-driven arousal ^25, 26^, it is anticipated that the observed pattern of reduced amygdala-orbitofrontal coupling primarily reflects a reduced need for top-down modulation. This interpretation is corroborated by the identified relationship between amygdala-orbitofrontal adaptations and changes in cardiac autonomic activity, predominantly in terms of enhanced parasympathetic high-frequency HRV. Importantly, this relationship indicates that children who display more pronounced attenuations in amygdala-orbitofrontal coupling, also display stronger oxytocin-induced increases in cardiac parasympathetic drive, i.e., reflective of a facilitation of the stress-regulatory ‘rest & digest’ autonomic state ^29^.

Previous acute effect studies yielded a pattern of oxytocin-induced increases in resting-state high-frequency HRV in healthy adults ^31, 32, 34^, patients with obstructive sleep apnea ^30^, individuals at risk for psychosis ^33^ and pregnant women ^48^ (although see ^49^). Note however, that studies examining oxytocin-induced changes in task-related HRV predominantly demonstrated an opposite pattern, indicating reduced cardio-parasympathetic tone, for instance during a mental arithmetic task in patients with chronic back pain ^50^ as well as during a mildly stressful social challenge in patients with Fragile-X-syndrome ^51^ (although see ^52^). This provides support to the notion that the impact of oxytocin may not be uniform and that the direction of effects may depend on the context in which oxytocin is administered. Accordingly, and as outlined above, it appears that particularly in ‘task-free’ conditions, oxytocin may further facilitate the body’s parasympathetic tone ^30-34, 48^. Upon mildly stressful tasks, on the other hand, oxytocin may primarily induce an increase in cardiac-sympathetic tone, likely reflective of enhanced attention and vigilance to the presented task ^50, 51^, as proposed in the Social Salience Hypothesis of oxytocin ^53^. Also with regard to chronic oxytocin administration studies, consensus is growing that variable treatment responses may at least be partly attributed to the fact that, in contrast to acute dosing studies, there is often a lack of standardization of the context in which the daily oxytocin nasal spray is administered. In a recent combinatory trial, significant clinical improvements in social functioning were observed in young children with ASD receiving oxytocin in combination with standardized parent-child social interactions, held within the two hours after the daily nasal spray administrations ^15^. These and other observations ^19, 54^ therefore prompt future trials to administer the daily doses of chronic oxytocin administrations within more standardized contexts, e.g. by pairing them with psychosocial trainings.

Aside context, also distinct person-dependent factors are put forward to contribute to the observed variability in treatment responses within and across studies. In line with this notion, we here showed that oxytocin’s neural and cardiac autonomic effects were significantly modulated by epigenetic variations of the *OXTR*, such that oxytocin-induced changes were more pronounced in children with lower levels of *OXTR* methylation (associated with higher oxytocin receptor expression). Albeit interpretative, these observations provide indications that chronic oxytocin administration may be predominantly beneficial for children who display at least a minimal level/availability of *OXTR* expression. While, to our knowledge, this is the first oxytocin trial to examine *OXTR* methylation as a potential treatment modulator, prior work in patients with obsessive compulsive disorder (OCD) has proposed *OXTR* DNA methylation to constitute an important biomarker of cognitive behavioral therapy (CBT) treatment responses ^55, 56^. Particularly, two recent trials consistently demonstrated *OXTR* hypermethylation (lower receptor expression) to be associated with impaired CBT treatment responses in OCD patients 55, 56.

With regard to diagnosis-related alterations in *OXTR* DNA methylation in ASD, evidence is currently sparse and inconclusive. A recent review pointed towards differential ASD-related alterations in *OXTR* DNA methylation depending on developmental stage suggesting a predominant pattern of hypomethylation in children, but hypermethylation in adults with ASD ^46^. The number of existing studies is limited, however, and overall sample sizes were small to modest. Also considerable design-related variations can be noted (e.g. in terms of included CpG sites) rendering it difficult to draw conclusive interpretations regarding diagnosis-specific alterations in *OXTR* DNA methylation in ASD ^46^. Despite these difficulties in delineating consistent categorical diagnosis-related alterations, the current research provides important indications that dimensional variations among ASD individuals may form an informative biomarker for delineating subgroups of patients that may benefit the most from chronic oxytocin administration.

Exploratory analyses of oxytocin-induced effects on whole-brain amygdala connectivity showed remarkable similarities between those reported in adults with ASD ^27^ and those seen in the current study in children with ASD. Particularly, both in adults and in the current pediatric population, an overall pattern of attenuated amygdala coupling was evident, indicating reduced functional connectivity to distinct regions including orbitofrontal cortex, as well as fusiform cortex, brainstem, opercular cortex, globus pallidus, supramarginal gyrus, and (para)hippocampal regions. Notably however, while in adults with ASD, this broader pattern of attenuated amygdala connectivity already emerged immediately post-treatment ^27^, it appeared that in children with ASD, a different time course of oxytocin’s amygdala effects was evident, with attenuating effects only significantly emerging at the T2 follow-up session, four weeks after cessation of the last nasal spray administration. Immediately after the nasal spray administration period (T1) on the other hand, children with ASD displayed a predominant pattern of enhanced amygdala coupling to distinct brain regions, particularly to regions of the salience network, including bilateral insular cortex, the anterior cingulate cortex and supplementary motor area. While speculative, the divergent pattern of neural amygdala effects in children, compared to adults immediately post-treatment may reflect a differential mechanistic recruitment of neural circuits. Particularly, in children the enhanced amygdala connectivity to the salience network may indicate a stronger reliance on oxytocin’s (social) salience enhancing effects ^53^, likely facilitating increased attention attribution to and awareness of sensory, emotional and cognitive information ^57^. In adults with ASD, on the other hand, there appeared to be a stronger immediate reliance on oxytocin’s stress-regulatory, anxiolytic role as reflected by the overall pattern of attenuated amygdala connectivity e.g. to orbitofrontal cortex and brainstem ^24, 27^. In children with ASD, these anxiolytic, amygdala attenuating effects appear to emerge only at a later stage, rendering oxytocin’s long-lasting neuromodulatory changes in children to be similar to those observed in adults. Future research will be needed to gain deeper insights into these divergent results patterns, and to discern whether potential differences in dosing schema (one daily dose of 24 IU in the adults, versus two daily doses of 12 IU in the pediatric sample) and/or differences in the duration of acute dosing effects may constitute variability inducing factors. For example, in adult populations, heightened levels of circulating oxytocin were shown up to 7 hours after acute administration ^58^. It is however unclear whether the duration of acute dosing effects is similar in pediatric populations or whether they can endure longer, potentially rendering the reported T1 post effect (assessed 24h after the last nasal spray) to be susceptible of reflecting – at least in part – an acute oxytocin dosing effect, rather than solely reflecting oxytocin’s recursive, chronic action on amygdala-centered circuits.

While the current study provides important new insights into the neural and cardiac autonomic effects of chronic oxytocin administration in children with ASD, the following limitations and recommendations are noted. First, a rather homogenous group of high-functioning children within a tight pre-pubertal age range were included, rendering generalizability of the identified oxytocin-induced effects to more heterogeneous or younger/older children and adolescents uncertain. Also, while the included number of boys and girls in our sample reflected the well-documented four-to-one male bias in ASD prevalence ^59^, future research is warranted to examine the observed oxytocin-induced effects also in larger samples of girls with ASD, especially given prior reports of sex-related differences in neural and behavioral responses to oxytocin ^60^. Finally, in the current study, participants administered 12 IU of the oxytocin nasal spray twice a day in the morning and afternoon, similar to ^13^, although considerable variation in dosing schemes can be noted among prior pediatric trials, e.g. one daily dose of 24 IU ^14, 17^ or intermittent ^15^ and flexible daily dosings ranging from 8 IU to 80 IU ^18^. Future trials should therefore be directed at identifying optimal daily dosing schemes, administration length and intervals.

To conclude, in this pharmaco-neuroimaging study in a pediatric population of children with ASD, we demonstrated important stress-regulatory neural and cardiac effects of chronic oxytocin administration, indicative of a facilitation of the ‘rest and digest’ parasympathetic autonomic state, even up to four weeks after cessation of the nasal spray administration period. Further, epigenetic profiling showed that inter-individual variation in *OXTR* DNA methylation may form an important biomarker for delineating subgroups of children with ASD who may benefit the most from a chronic regime of oxytocin administrations.

## Data Availability

All data produced in the present study are available upon reasonable request to the authors

## Acknowledgements

We would like to thank all the participants of the study and our colleagues of the Leuven Autism Research Consortium (LAuRes).

This research was supported by an internal C1 and Small Research Equipment fund of the KU Leuven (ELG-D2857-C14/17/102; KA/20/080), a Doctor Gustave Delport fund of the King Baudouin Foundation, the Branco Weiss fellowship of the Society in Science - ETH Zurich, an Excellence of Science EOS grant (G0E8718N) granted to KA or BB. JP is supported by the Marguerite-Marie Delacroix foundation and a postdoctoral fellowship of the Flanders Fund for Scientific Research (FWO; 1257621N). SVDD is supported by a FWO postdoctoral fellowship (12C9723N). MM is supported by a KUL postdoctoral mandate. ME is supported by an FWO aspirant fundamental fellowship (11N1222N).

The funding sources had no further role in study design; in the collection, analysis and interpretation of data; in the writing of the report; and in the decision to submit the paper for publication.

## Statement of Ethics

This study protocol was reviewed and approved by the Ethics Committee for Biomedical Research at the University of Leuven, approval number [S61358]. Written informed consent from the parents and assent from the child were obtained prior to the study.

## Supplementary Material

### Supplementary Methods

#### Participants and compliance/ side effects monitoring

##### COVID-19 related impact on recruitment

As indicated in the initial EudraCT trial registration, fMRI data collections were part of a broader assessment including behavioral-clinical characterizations ^1^, as well as other (neuro)physiological (electroencephalographic) and biological assessments, for which a total sample size of 60 participants was planned (30 in each treatment arm). However, due to COVID-19 related health restrictions, all physiological, neural and neurophysiological data collections were temporarily halted, resulting in extensive loss of follow-up data on these neural outcomes. To account for this data loss, and upon approval from the Ethical committee, recruitment was extended after the COVID-19 lock-downs, to further increase the total sample size of the neural and neurophysiological assessments. Accordingly, the total number of randomized participants in the trial was 80 participants (40 oxytocin, 40 placebo) as visualized in the CONSORT flow diagram in **Figure 1**.

##### Compliance monitoring

Compliance was assured using a daily medication diary that recorded date and time of administration (percentage compliance; oxytocin: 96.75 ± 5.26%; placebo: 96.11 ± 5.29 %; *t*(74) = .52, *p* = .603. The total amount of administered fluid was also monitored (oxytocin: 14.86 ± 2.37 ml; PL: 13.79 ± 2.35 ml; *t*(75) = 2.00, *p* = .05).

##### Side effects

During the course of the nasal spray administration period, participants were screened for potential adverse events (weekly parent report) or changes in affect and arousal (daily diary by child and parent). Overall, reports of side effects were minimal and not specific to the oxytocin nasal spray (see ^1^ and adverse event reporting at the EU Clinical Trials register - EudraCT 2018-000769-35).

#### fMRI scanning

##### MRI data scanning parameters

Anatomical imaging consisted of a high-resolution structural volume acquired using a coronal three-dimensional turbo field echo T1-weighted sequence with the following parameters: 160 contiguous sagittal slices covering the whole brain and brainstem, slice thickness = 1.2 mm; repetition time (TR) = 9.6 ms; echo time (TE) = 4.6 ms; matrix size = 256 × 256; field-of-view (FOV) = 250 × 250 mm; in-plane pixel size = 0.98 × 0.98 mm^2^; acquisition time = 5 min 37.8 s.

Resting-state fMRI images were acquired using a whole brain multiband T2*-weighted gradient-echo echo planar imaging (GE-EPI) sequence with the following parameters: TR = 1500 ms; TE = 30 ms; matrix size = 84 × 82, FOV = 228 × 228 mm; flip angle 80°; slice thickness = 2.75 mm, slice gap = 0 mm; multi-band factor = 2; axial slices = 48; 270 functional volumes; acquisition time = 6 min 57 s. During the resting-state scan, participants were asked to stare at a white cross against a black background, relax, lie still and to not think of anything in particular.

Note that the fMRI scanning protocol additionally included two other scan modalities: (i) task-based fMRI scanning and (ii) diffusion tensor imaging at baseline. The analyses of these scan modalities are not part of the current report.

##### MRI data pre-processing

Pre-processing analyses of the raw functional MRI data were performed using the Functional Connectivity Toolbox (CONN version 17.f) ^2^, implemented in Matlab R2020b (Mathworks).

Resting-state fMRI images were slice-time corrected, spatially realigned, normalized to the standard EPI-template of the Montreal Neurological Institute (MNI-152) and resampled into 2-mm isotropic voxels. Further, the data were spatially smoothed with a Gaussian kernel of 5 mm full width half maximum (FWHM) ^3^. Realignment parameters were modelled as regressors of no-interest and white matter and cerebrospinal fluid were removed as confounds following the implemented aCompCor-strategy in the CONN toolbox ^4^. Residual time-series of the resting-state images were then band-pass filtered (0.008 Hz<f<0.09 Hz).

##### Head motion

Given the potential confounding effects of head micromovements on resting-state functional connectivity, all reported analyses were performed on ‘scrubbed’ data ^5^, i.e., censoring frames displaying frame-wise displacement exceeding > 0.9 mm or frame-wise changes in brain image intensity exceeding > 5 SD. On average, approximately 9.9% of the frames were censored per subject. No nasal spray (oxytocin, placebo) (*F*(1,53.35) = 1.49; *p* = .22; *ŋ*^*2*^ = .03) or session-related (T0, T1, T2) differences (*F*(2,115.74) = 0.26; *p* = .77; *ŋ*^*2*^ = .004) were revealed in the number of scrubbed frames. Also, secondary analyses including the number of scrubbed frames or mean motion as a ‘nuisance’ covariate did not qualitatively change the pattern of results obtained from the reported mixed-effects analyses.

#### Assessment of baseline DNA methylation of the oxytocin receptor gene

##### Sample collection

Salivary samples were collected from each child at the end of their baseline (T0) study visit using the Oragene DNA sample collection kit (DNA Genotek Inc., Canada), to explore epigenetic variations of the oxytocin receptor gene (*OXTR)*. Salivary collections constitute a fairly simple and stress-free method that is easily applicable in vulnerable (pediatric) populations. Furthermore, recent evidence indicates that it is as reliable and suitable for DNAm analyses as methylation patterns detected in other tissues such as brain and blood ^6^. Oragene DNA saliva samples were collected in the afternoon, on average at 15:36h (range 11:50 – 19:07).

##### Sample analysis

After data collection, 200 ng DNA was extracted from the samples and bisulfite converted following the manufacturer’s protocol (EZ-96 DNAm Kit, Zymo Research, Irvine, CA, USA). Bisulfite converted DNA was stored at -80 °C until further analysis. Next, the levels of methylation at three CpG sites (i.e., -934, -924 and -914) of *OXTR* (hg19, chr3:8,810,729-8,810,845) were determined using Pyrosequencer (Qiagen, Hilden, Germany) and analyzed using Pyromark Q96 software. Laboratory procedures and analyses were conducted in accordance with manufacturer’s protocols and software settings ^7^. Protocols for the PCR amplification and Pyrosequencing analysis were adapted from ^8^.

##### PCR primers

The following PCR primers were adopted to amplify the DNA fragment of interest at the CpG sites -934, -924 and -914: [OXTR Forward: TTG AGT TTT GGA TTT AGA TAA TTA AGG ATT; OXTR Reverse: /5Biosg/AC TTA ACA TCA CAT TAA ATA CAA CC]. The PCR conditions during the amplification were as follows: step 1: (95°C/15 min)/1 cycle; step 2: (94°C/30 s, 58°C/30 s, 72°C/30 s)/50 cycles; step 3: (72°C/10 min)/1 cycle; and step 4: 4°C hold. Also the following sequencing primer was used to read the DNA sequence (OXTR Sequencing: AGA AGT TAT TTT ATA ATT TT).

**Supplementary Table 1.**
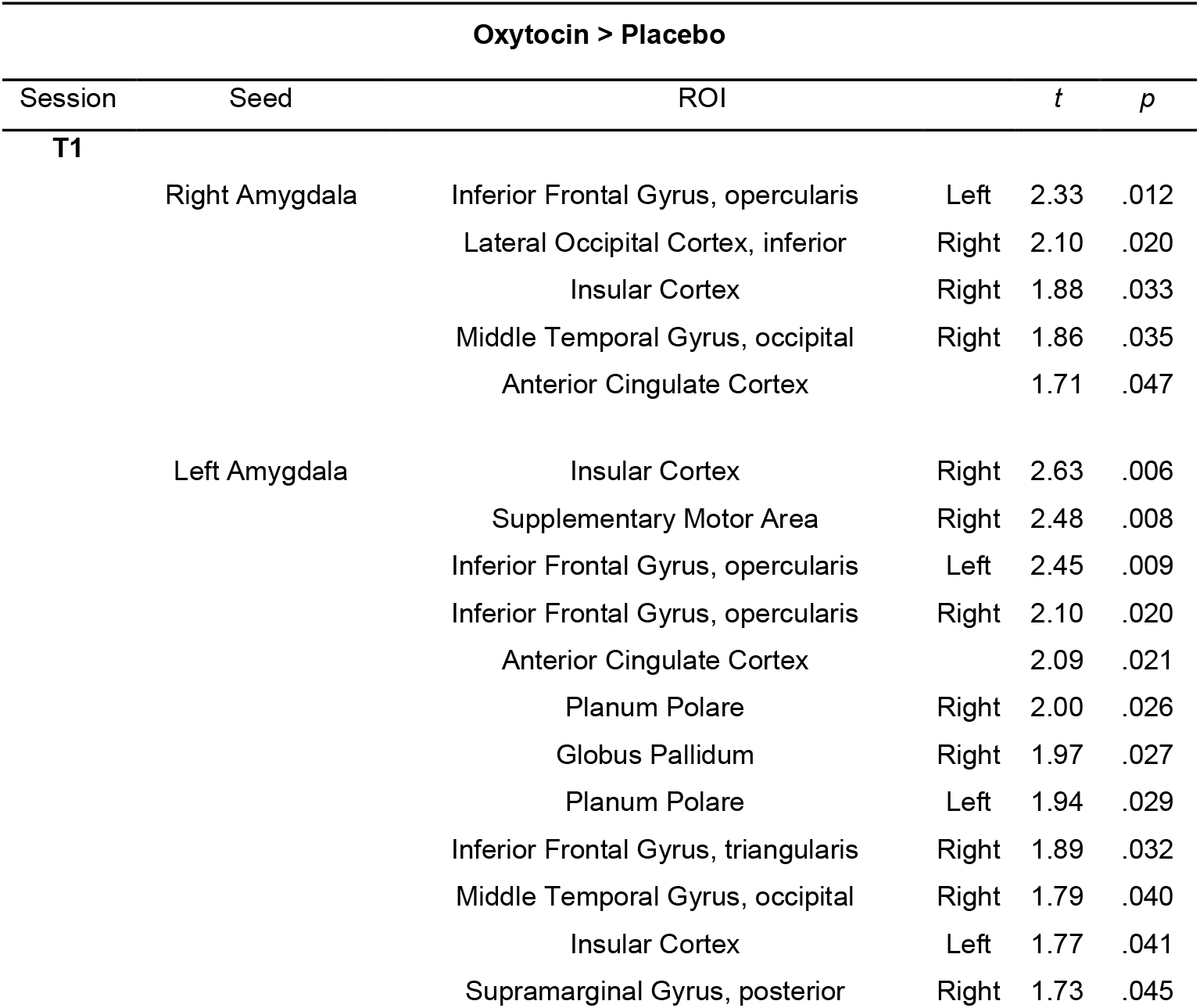
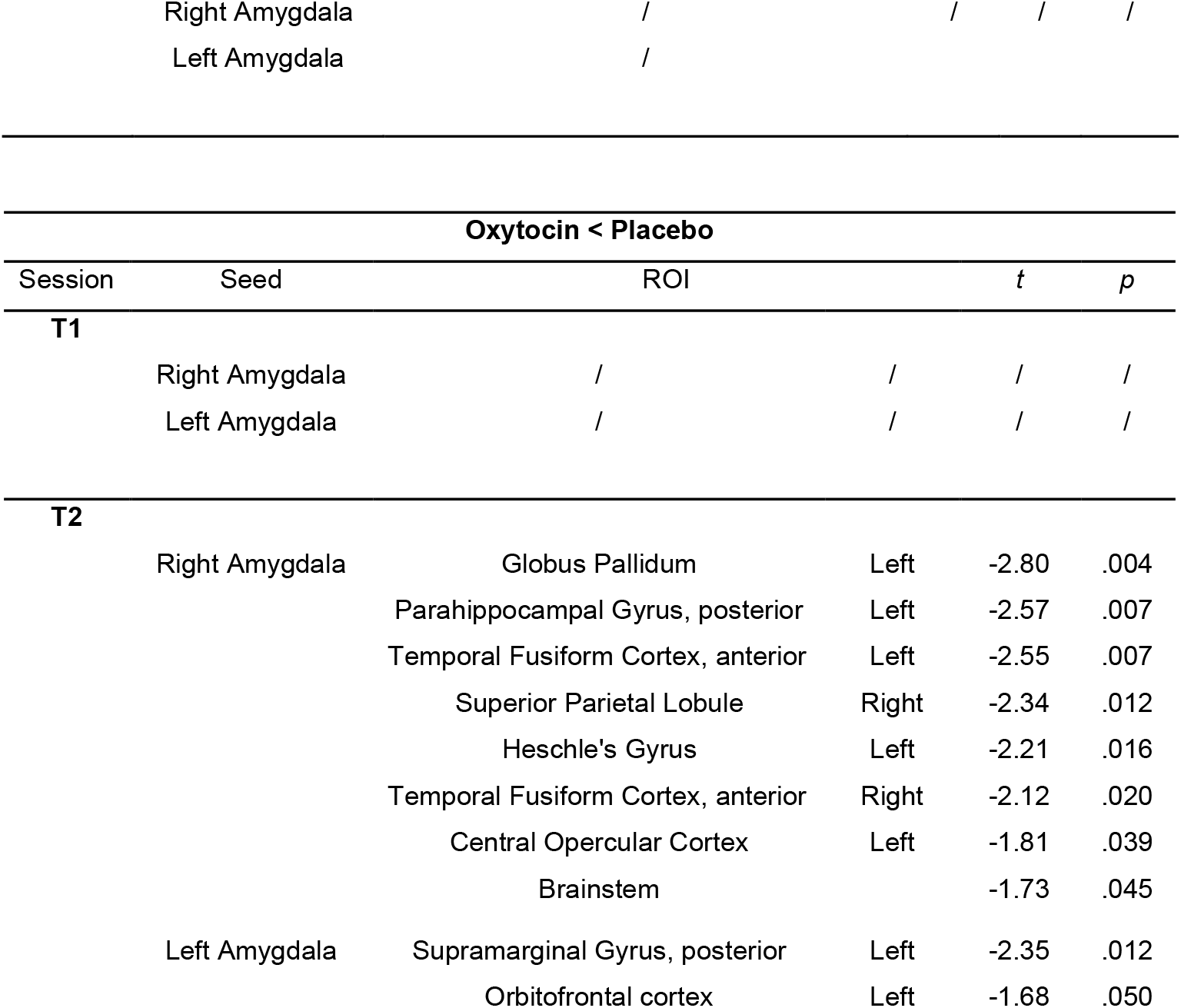
Oxytocin-induced changes in whole-brain amygdala connectivity. Exploratory whole-brain analyses were performed by assessing oxytocin-induced changes in functional connectivity between bilateral amygdala (seeds) and all other regions of the cortical (*n* = 91) and subcortical (*n* = 13) Harvard-Oxford atlas. At the **T1 post session**, an overall pattern of enhanced amygdala connectivity was identified, particularly to regions of the salience network (e.g., bilateral insular cortex, posterior insula in planum polare, anterior cingulate cortex, right supplementary motor area). At the **T2 follow-up session**, an overall pattern of reduced amygdala connectivity was evident, to distinct cortical and subcortical regions. Connections displaying significant oxytocin-induced effects (independent *t*-test) are reported separately for each assessment session (T1, T2) at an uncorrected *p* < .05 threshold.

